# Blood transcriptomics reveal persistent SARS-CoV-2 RNA and candidate biomarkers in Long COVID patients

**DOI:** 10.1101/2024.01.14.24301293

**Authors:** Soraya Maria Menezes, Marc Jamoulle, Maria Paula Carletto, Leen Moens, Isabelle Meyts, Piet Maes, Johan Van Weyenbergh

## Abstract

With an estimated 65 million individuals suffering from Long COVID, validated therapeutic strategies as well as non-invasive biomarkers are direly needed to guide clinical management. We used blood digital transcriptomics in search of viral persistence and Long COVID diagnostic biomarkers in a real-world, general practice-based setting with a long clinical follow-up. We demonstrate systemic SARS-CoV-2 persistence for more than 2 years after acute COVID-19 infection. A 2-gene biomarker, including *FYN* and SARS-CoV-2 antisense RNA, correctly classifies Long COVID with 93.8% sensitivity and 91.7% specificity. Specific immune transcripts and immunometabolism score correlate to systemic viral load and patient-reported anxiety/depression, providing mechanistic links as well as therapeutic targets to tackle Long COVID.

## Introduction

With an estimated 65 million individuals suffering from Long COVID^1^, validated therapeutic strategies as well as non-invasive biomarkers are direly needed to guide clinical management. We used blood transcriptomics in search of viral persistence and Long COVID diagnostic biomarkers in a real-world, general practice-based setting with a long clinical follow-up (median 2 years).

## Methods

Long COVID patients were diagnosed according to WHO criteria and followed up for up to 39 months after acute COVID^2^. Complete clinical history was obtained using electronic health records and validated clinical scales (Duke Severity of Illness, Dartmouth Coop charts, see Supplementary Data) were used to quantify patient evolution^2^. Whole blood samples were obtained from 48 Long COVID patients and 12 controls from the same general practice, matched for age, sex, time since acute COVID-19 and severity (47/48 patients and 12/12 mild-moderate, non-hospitalized), vaccination status and comorbidities (Supplementary Table 1) and analyzed by digital transcriptomic analysis (nCounter, Nanostring), as previously established for critical COVID-19^3^. Differentially expressed genes and predefined biological pathway scores were determined using nSolver (detailed gene lists in Supplementary Table 2). Correction for multiple testing was performed by the Benjamini-Hochberg method with a False Discovery Rate (FDR) cut-off of 5%. Total blood viral load was determined as the sum of all individual SARS-CoV-2 transcripts. Statistical tests performed with GraphPad Prism and XL-STAT software included normality testing (Shapiro-Wilk and Kolmogorov-Smirnov tests), which guided subsequent parametric (t-test) or non-parametric (Mann-Whitney test, Spearman correlation) analysis, all two-tailed.

## Results

Digital transcriptomic analysis showed a total of 212 differentially expressed genes (uncorrected p<0.05) between Long COVID patients and matched controls (Fig. 1A), of which 70 genes remained significant after FDR correction (Suppl. Table 2). Among the up-regulated transcripts were several viral RNAs: Nucleocapsid, ORF7a, ORF3a, Mpro (target of Paxlovid) and antisense ORF1ab RNA, the latter suggesting ongoing viral replication, while Spike RNA was low. In addition, several SARSCoV2-related host genes were also increased in Long COVID (*ACE2/TMPRSS2* (co)receptors and *DPP4/FURIN* proteases). Other upregulated RNAs were prototypic for memory B cells and platelets (Fig. 1A). ROC curve analysis shows significant discrimination (AUC 0.94 95% CI [0.86-1.00], p=3×10^-6^) between Long COVID patients (n=48) and matched controls (n=12), with 93.8% sensitivity and 91.7% specificity (Fig. 1B). Multivariable logistic regression showed antisense SARS-CoV-2 and *FYN* RNA levels were independent predictors of disease status (corrected for age and sex, see Supplementary Table 2). As single biomarkers, antisense SARS-CoV-2 (AUC 0.78 95% CI [0.65-0.90], p=0.0033) and *FYN* RNA (AUC 0.89 95% CI [0.79-0.99], p=3×10^-5^) were also significant predictors of Long COVID disease status, but with lower sensitivity and specificity (Fig. 1B). Summarizing transcriptomic results into biological pathways, we found significantly decreased lymphocyte activation (p=0.016) and immunometabolism (p=0.023) in Long COVID patients (Fig. 1C). Moreover, immunometabolism score was negatively correlated with total blood viral load (Fig. 1D, R=-0.56, p<0.0001).

**Figure 1.**
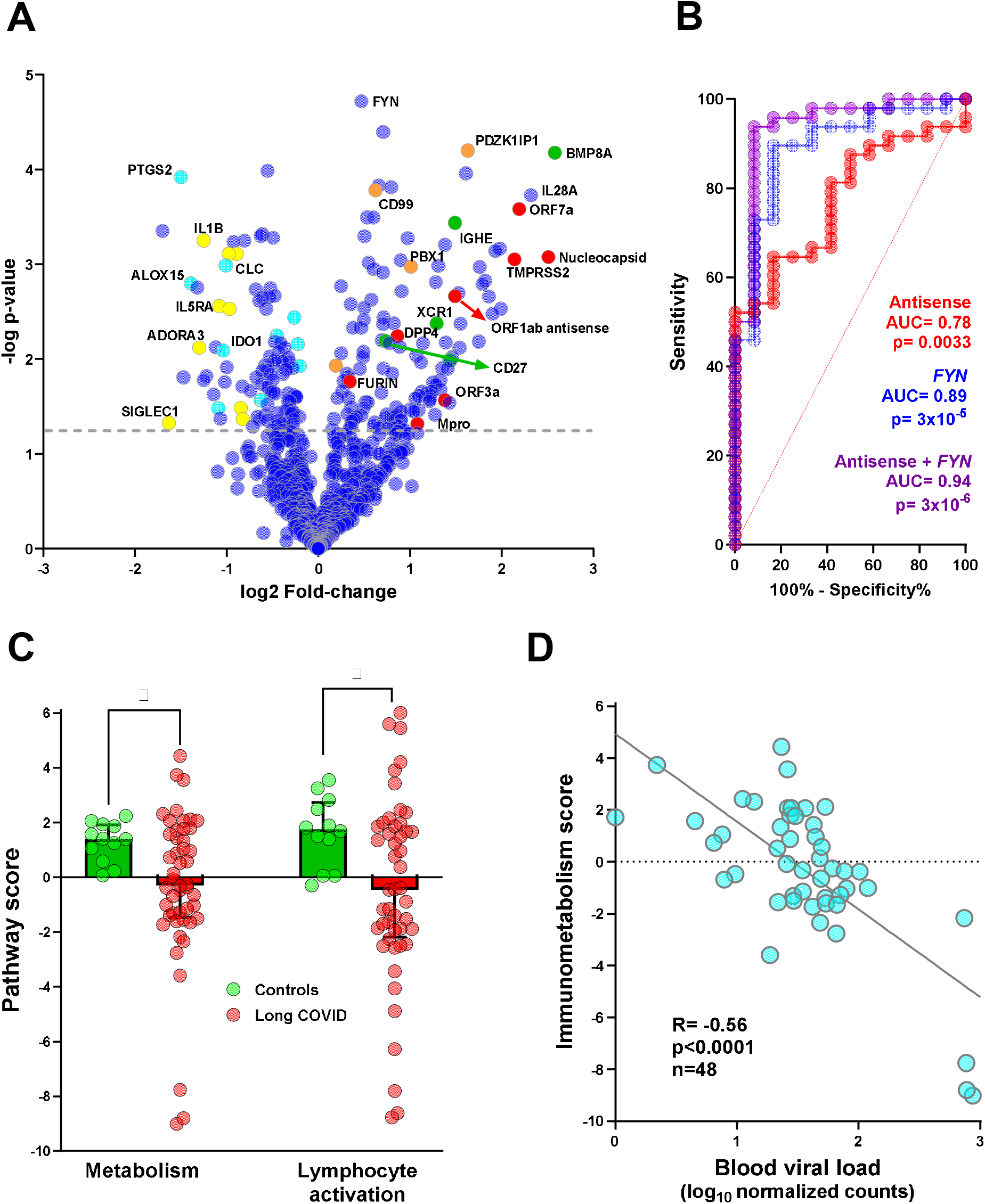
A) Volcano plot of differentially expressed genes in whole blood samples between Long COVID patients (n=48) and controls (n=12) matched for age, sex, vaccine status, time since acute COVID-19 and number of comorbidities (Supplementary Table 1). Digital transcriptomics (nCounter, Nanostring Technologies Ltd.) was used as previously described^3^. Genes highlighted in red correspond to viral RNAs (Nucleocapsid, ORF3a, ORF7a, Mpro and antisense ORF1ab) and SARS-CoV-2-related host transcripts (*ACE2/TMPRSS2* (co)receptors, *DPP4* and *FURIN* proteases). Genes highlighted in green and salmon correspond to memory B-cell (*BMP8A, IGHE, CD27, XCR1*) and platelet-expressed (*PDZK1IP1, PBX1, CD99*) transcripts, respectively. Genes highlighted in turquoise and yellow represent transcripts belonging to immunometabolism (*PTGS2, ALOX15, IDO1* and others) and lymphocyte activation (*IL5RA, ADORA3A, SIGLEC1, IL1B* and others) biological pathways, respectively (detailed in Supplementary Data). B) ROC curve analysis shows significant discrimination (AUC 0.94 95% CI [0.86-1.00], p=3×10^-6^) between Long COVID patients (n=48) and matched controls (n=12), as determined by multivariable logistic regression with antisense SARS-CoV-2 and *FYN* transcript levels as independent predictors (corrected for age and sex, see Supplementary Table 2). As single biomarkers, antisense SARS-CoV-2 (AUC 0.76 95% CI [0.86-1.00], p=3×10^-6^) and FYN RNA (AUC 95% CI [0.86-1.00], p=3×10^-6^) were also significant predictors of Long COVID disease status, but with lower sensitivity and specificity. C) Significant decrease in immunometabolism and lymphocyte activation scores (quantified by nCounter digital transcriptomics) in Long COVID patients (n=48) compared to matched controls (n=12), bars represent the median with 95% CI (Mann-Whitney test *p<0.05) D) Negative correlation between immunometabolism score and viral load (sum of all SARS-CoV-2 transcripts detectable above background), as quantified by digital transcriptomics (Spearman correlation, n=48).

In addition to quantitative analysis (Fig. 1A), we also performed qualitative analysis of each SARS-CoV-2 transcript as well as total blood viral load, comparing the frequency of positive (above cut-off, Fig. 2A) and negative individuals (below cut-off) in each group. Significant differences between Long COVID patients and matched controls were observed for SARS-CoV-2 Antisense (65% vs. 25% positives, respectively, p<0.05) , ORF7a (60% vs. 25% positives, respectively, p<0.05) and N (Nucleocapsid, 50% vs. 8% positives, respectively, p<0.01) RNAs, as well as total blood viral load (60% vs. 8% positives, respectively p<0.01). Due to the large variation in blood viral load in Long COVID patients (Fig. 2A), we used multivariable regression to find demographic or clinical predictors of low’ vs. ‘high’ viral load status (above or below cut-off, respectively). We found that age and sex were not associated with ‘low’ vs. ‘high’ viral RNA status, whereas the number of comorbidities (1.61 95% CI [1.14-2.49], p=0.014) and the number of COVID vaccine doses (0.36 95% CI [0.14-0.79], p=0.018) were independent predictors of ‘low’ vs. ‘high’ status (Supplementary Table 3). In addition, we observed a highly significant positive correlation between immune/platelet transcripts (*PDZK1IP1, CD99*) and total blood viral load (Fig. 2B).

**Figure 2.**
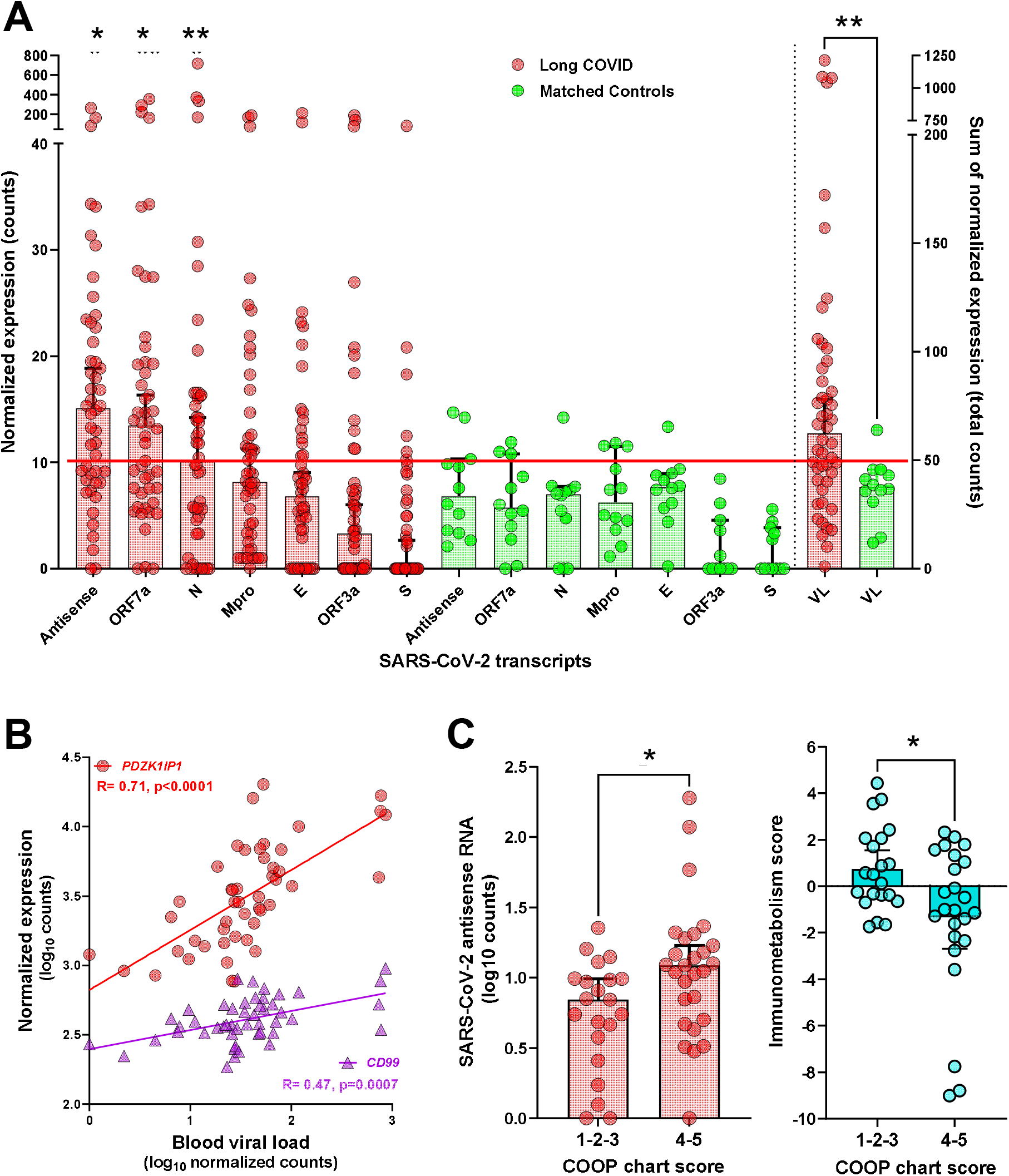
A) Overview of individual data for all SARS-CoV-2 transcripts (normalized expression in counts) and total blood viral load (sum of all SARS-CoV-2 normalized counts). Each circle represents a single Long COVID patient (red, n=48) or a matched control (green, n=12), bars represent the median with 95% CI. The red horizontal lane represents the cut-off for positivity (10 normalized counts for individual transcripts and 50 normalized counts for total viral load). Significant differences between Long COVID patients and controls were determined by Fisher’s test (*p<0.05, **p<0.01). B) Positive correlation between immune/platelet transcripts (*PDZK1IP1, CD99*) and viral load (sum of all SARS-CoV-2 transcripts detectable above background), as quantified by digital transcriptomics (Spearman correlation, n=48). C) Patient-reported outcome measures (COOP chart score on the question “During the last two weeks, how much have you been bothered by emotional problems such as feeling anxious, depressed, irritable, or downhearted and sad?”, on a visual scale from 1 to 5, with 5 being worst). As compared to patients grouped as ‘mild’ (score1-2-3, n=21), ‘severe’ patients (score 4-5, n=23) were significantly associated with higher SARS-CoV-2 antisense RNA levels (left panel, *p<0.05 Unpaired t test) and lower immunometabolism score (right panel, *p<0.05 Mann-Whitney test). Bars represent the median and 95% CI.

Finally, we found that viral RNA and immunometabolism score were linked to patient-reported outcome measures (COOP chart emotional ‘anxiety/depression’ score, see Suppl. Figure 1). As compared to patients grouped as ‘mild’ (score1-2-3, n=21), ‘severe’ patients (score 4-5, n=23) were significantly associated with higher SARS-CoV-2 antisense RNA levels (left panel, *p<0.05 Unpaired t test) and lower immunometabolism score (right panel, *p<0.05 Mann-Whitney test).

## Discussion

We demonstrate SARS-CoV-2 viral RNA persistence in Long COVID patients compared to matched post-pandemic controls, at higher frequencies than recently reported at the protein level^4^ (max. difference 52% vs. 11%) and for a longer period after acute COVID-19 infection (>24 months vs. 10-14 months). To our knowledge, this study provides the first blood transcriptome Long COVID biomarker with >90% sensitivity and specificity, hence amenable for large-scale diagnostic testing on the robust nCounter platform. This candidate diagnostic 2-gene blood biomarker, identified in a real-world setting, remains to be validated in independent Long COVID cohorts.

Of note, platelet-expressed transcripts were positively correlated to viral load (Fig. 2B), providing a mechanistic link to the hypercoagulative state previously demonstrated in Long COVID^1^, as well as a possible viral reservoir^5^. On the other hand, immunometabolism score was negatively correlated with blood viral load, suggesting a decreased metabolic status in Long COVID due to ongoing viral replication. In support of this hypothesis, patient-reported outcome measures (COOP chart emotional scores) were significantly associated with both SARS-CoV-2 antisense RNA, a surrogate marker of viral replication, and immunometabolism score (Fig. 2C).

In conclusion, blood transcriptomics reveal systemic SARS-CoV-2 persistence up to more than 2 years after acute COVID-19 infection. Specific immune transcripts and immunometabolism score correlate to systemic viral load and patient-reported anxiety/depression, providing mechanistic links as well as therapeutic targets to tackle Long COVID.

## Data Availability

All data produced in the present study are available upon reasonable request to the authors.

