## Supplementary Data for "Blood transcriptomics reveal persistent SARS-CoV-2 RNA and candidate biomarkers in Long COVID patients"

**Supplementary Data to Menezes et al.**

This study was approved by the Ethical committee of University Hospitals Leuven (UZ Leuven), all patients and controls have provided written informed consent.

Whole blood samples were obtained from 48 Long COVID patients and 12 controls matched for age, sex, time since acute COVID-19, vaccination status and comorbidities, as shown in Supplementary Table 1. A diagnosis of Long COVID was made according to WHO criteria^1^: “defined as the continuation or development of new symptoms 3 months after the initial SARS-CoV-2 infection, with these symptoms lasting for at least 2 months with no other explanation.” Patient-reported outcomes were measured using validated COOP/WONCA charts^2^ (see Supplementary Figure 1), clinician-reported outcomes were measured using DUSOI (Duke University Severity Of Illness) score^3^.

Acute COVID-19 infection and comorbidities were documented for all patients and controls using the national database of electronic health records, which is centralized and linked to a single general practician per patient in the Belgian healthcare system (access granted to co-author MJ for all patients and controls).

Host and viral gene expression was quantified simultaneously in a single blood sample by digital transcriptomic analysis (nCounter, Nanostring Technologies), as we previously established in critical COVID-19^4^, fatal COVID-19 nursing home outbreaks^5^, respiratory infections in the Emergency department^6^, and other viral infections^7-8^. In short, >800 transcripts corresponding to host immune (Myeloid and Innate Immunity panel) including 40 housekeeping genes, as well as SARS-CoV-2 viral transcripts and *ACE2/TMPRSS2* receptors (customized Plus Panel), were quantified by hybridization with specific fluorescent barcodes linked to a 50 bp reporter probe and an adjacent 50 bp capture probe, followed by normalization for background (negative control probes), internal positive control probes and housekeeping genes using nSolver software. Differentially expressed genes and predefined biological pathway scores were determined using nSolver (detailed gene lists in Supplementary Table 2). Statistical tests performed with GraphPad Prism and XL-STAT software included normality testing (Shapiro-Wilk and Kolmogorov-Smirnov tests), which guided subsequent parametric (t-test) or non-parametric (Mann-Whitney test, Spearman correlation) analysis, all two-tailed.

**Disclosure of all financial associations or other possible conflicts of interest:**

None of the authors have a potential conflict of interest.

**Supplementary Table 1: Clinical and demographic data for Long COVID patients and controls**

|  | **Patients** | **Controls** |
| --- | --- | --- |
| **Age (years)** | 43.0 (IQR 32.3-54.8) | 47.0 (IQR 34.3-64.5) |
| **Sex** | 15 M/33 F (68%) | 4M /8F (67%) |
| **Number of comorbidities** | 3 (IQR 2-5) | 2 (IQR 1-5) |
| **Number of vaccine doses** | 2 (IQR 2-3) | 3 (IQR 2-4) |
| **Time since acute COVID (months)** | 24 (IQR 15-32) | 21 (IQR 14-29) |
| **Acute COVID mild/moderate** | 47/48 | 12/12 |
| **DUSOI score** | 4 (IQR 3-4) | NA |
| **COOP total score** | 22 (IQR 19.8-24) | NA |
| **SPECT positive** | 27/48 (56%) | NA |

*Data represent median and interquartile range (IQR). DUSOI score (clinician-reported) ranges from 1-5, COOP total score (patient-reported) ranges from 6-30, higher scores correspond to worse clinical status for both. No statistically significant differences were observed for age, sex, number of comorbidities, number of vaccine doses or time since acute COVID-19 (Mann-Whitney test for all except Fisher’s test for sex).*

**Supplementary Table 3: Multivariable logistic regression model for classification of Long COVID patients (n=48) vs. matched controls (n=12)**

| **Variable** | **Odds ratio** | **95% CI** | **P value** |
| --- | --- | --- | --- |
| **Age** | 0.99 | 0.908-1.078 | 0.81 |
| **Sex[M]** | 0.66 | 0.083-4.709 | 0.67 |
| **SARS-CoV-2 antisense RNA** | 1.23 | 1.043-1.587 | 0.042 |
| ***FYN* RNA** | 1.01 | 1.003-1.014 | 0.0047 |

**Supplementary Table 4: Multivariable logistic regression model for ‘high’ vs. ‘low’ blood viral load status among Long COVID patients (n=48)**

| **Variable** | **Odds ratio** | **95% CI** | **P value** |
| --- | --- | --- | --- |
| **Age** | 0.96 | 0.89-1.02 | 0.20 |
| **Sex[M]** | 0.61 | 0.084-3.46 | 0.59 |
| **Number of comorbidities** | 1.61 | 1.14-2.49 | 0.014 |
| **Number of vaccine doses** | 0.36 | 0.14-0.79 | 0.018 |

**Supplementary Table 2: List of differentially expressed immune viral transcripts, and predefined biological pathways quantified by nCounter digital transcriptomics (Myeloid/Innnate Immunity panel, Nanostring Technologies): supplied as Excel file**

**Supplementary Figure 1: Visual chart (COOP/WONCA) used to quantify patient-reported outcomes in six dimensions (physical fitness, feelings, daily activities, social activities, change in health, overall health), each on a visual scale from 1 to 5 (higher scores means worse status).**

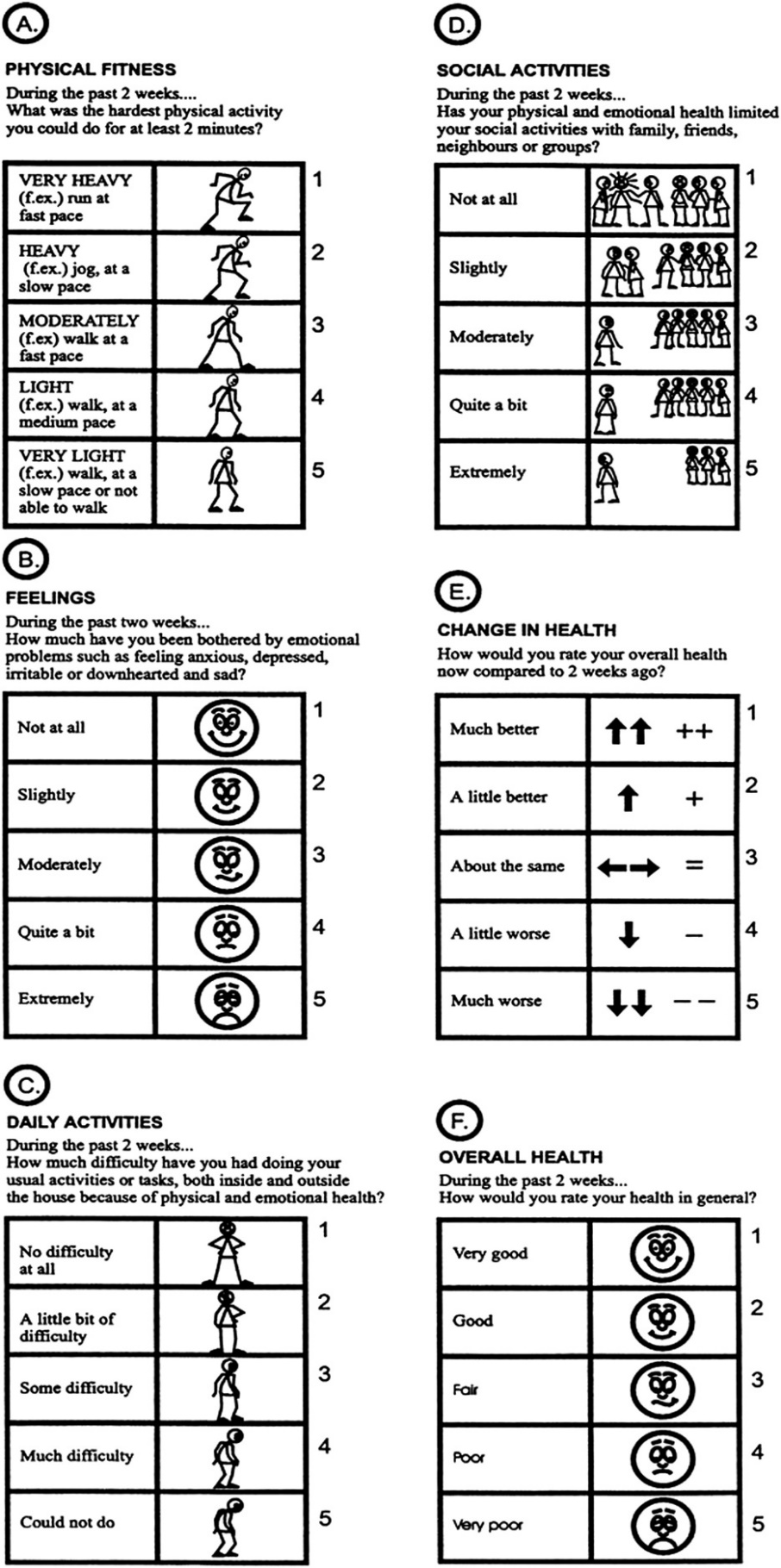
